# healthcareCOVID: A national cross-sectional observational study identifying risk factors for developing suspected or confirmed COVID-19 in UK healthcare workers

**DOI:** 10.1101/2020.08.28.20182295

**Authors:** J Kua, R Patel, E Nurmi, S Tian, H Gill, DJN Wong, C Moorley, D Nepogodiev, I Ahmad, K El-Boghdadly

## Abstract

**Objective:** To establish the prevalence and risk factors for the development of suspected or confirmed coronavirus disease 2019 (COVID-19) infection among healthcare workers (HCWs) in the United Kingdom (UK).

**Design:** Cross-sectional observational study.

**Setting:** UK-based primary and secondary care.

**Participants:** HCWs aged ≥18 years working between 1 February and 25 May 2020.

**Main outcome measures:** A composite endpoint of laboratory-confirmed diagnosis of SARS-CoV-2, or self-isolation or hospitalisation due to suspected or confirmed COVID-19.

**Results:** Of 6152 eligible responses, the composite endpoint was present in 1806 (29.4%) HCWs, of whom 49 (0.8%) were hospitalised, 459 (7.5%) tested positive for SARS-CoV-2, and 1776 (28.9%) reported self-isolation. The strongest risk factor associated with the presence of the primary composite endpoint was increasing frequency of contact with suspected or confirmed COVID-19 cases without adequate personal protective equipment (PPE): “Never” (reference), “Rarely” (adjusted odds ratio 1.06, (95% confidence interval: 0.87 to 1.29)), “Sometimes” (1.7 (1.37 to 2.10)), “Often” (1.84 (1.28 to 2.63)), “Always” (2.93, (1.75 to 5.06)). Additionally, several comorbidities (cancer, respiratory disease, and obesity); working in a ‘doctors’ role; using public transportation for work; regular contact with suspected or confirmed COVID-19 patients; and lack of PPE were also associated with the presence of the primary endpoint. 1382 (22.5%) HCWs reported lacking access to PPE items while having clinical contact with suspected or confirmed COVID-19 cases. Overall, between 11,870 and 21,158 days of self-isolation were required by the cohort, equalling approximately 71 to 127 working days lost per 1000 working days.

**Conclusions:** Suspected or confirmed COVID-19 was more common in HCWs than in the general population. Risk factors included inadequate PPE, which was reported by nearly a quarter of HCWs. Governments and policymakers must ensure adequate PPE is available as well as developing strategies to mitigate risk for high-risk HCWs during future COVID-19 waves.

## INTRODUCTION

Coronavirus disease 2019 (COVID-19), caused by severe acute respiratory syndrome coronavirus 2 (SARS-CoV-2), has resulted in a global health crisis that has challenged healthcare systems around the world. As of 09 August 2020, there were over 19 million confirmed cases and at least 721,000 deaths worldwide.^1^ Healthcare workers (HCWs) have been identified to be at risk of nosocomial COVID-19 infection.^2-6^ In the United States (US), over 124,000 HCWs have been infected with COVID-19,^7^ and up to 16% of those infected in the UK were thought to be key workers, a category that includes both HCWs as well as other essential workers from other industries.^8^

The prevalence of COVID-19 in HCWs is thought to be higher than in the general population due to exposure to higher viral loads from increased contact with infected individuals.^5,9,10^ The prevalence of COVID-19 amongst HCWs is variable with limited data comparing different staff roles and workplace environments (i.e. secondary versus primary care).^11-15^ Recent reports of HCW deaths have also revealed that Black, Asian and Minority Ethnic (BAME) groups appear to represent a greater proportion of these deaths.^3,4^ The reasons for this preponderance of BAME groups and COVID-19 severity in HCWs as well as the general population are likely to be complex and multifactorial.^18^ Similarly, HCW-related mortality from COVID-19 in the UK is reported as one of the highest globally, yet the reasons for this are poorly understood.^16^

Due to the nature of SARS-CoV-2 transmission, HCWs exposed to aerosol-generating procedures (AGPs) are at potentially higher risk of developing COVID-19.^5,17,18^ However, what constitutes an AGP remains contentious, with conflicting international and national guidance.^19,20^ Furthermore, shortages of personal protective equipment (PPE) throughout the pandemic and beyond remains a concern for HCWs.^21,22^ Taken together, the workplace environmental risks for HCWs with different exposures to COVID-19 and access to PPE remain unclear.

We therefore designed a UK-wide cross-sectional study to understand the prevalence and possible risk factors for the reporting of suspected or confirmed COVID-19 infection amongst HCWs. We aimed to capture details on socio-demographics, occupational exposure, and use of PPE to help expand the evidence base for HCWs and policymakers.

## METHODS

We conducted a cross-sectional observational study of UK-based HCWs between 4-25 May 2020 in England, Northern Ireland, Scotland and Wales. We included all HCWs aged 18 years or above and working at any time since 1 February 2020. Healthcare workers practicing in both primary (community and social care facilities) as well as secondary (hospitals) care were eligible. The study was prospectively registered as a service evaluation project at Guy’s and St Thomas’ NHS Foundation Trust (Service Evaluation ID: 10834) and was deemed to not require ethical approval by the Research and Development Department and the Health Research Authority Decision Tool.

### Study design

We designed an online survey using Knack (Evenly Odd Inc., Philadelphia, USA), an online data capture and database system. A multi-phased process involving several authors (JK, RP, KE, IA, DW, CM) was used to construct, revise and ratify the final survey. An initial draft of questions for the survey was created by JK and RP and sent to the remaining authors for review. Based on feedback received, modifications were made and questions compiled, followed by a second round of review and testing by all authors on the online system. This version of the survey was piloted in a convenience sample of 93 participants. One change was made as a consequence: an expanded list of specialties was implemented.

The final survey comprised 33 closed questions and five free-text entries, divided into five sections: (1) participant characteristics; (2) work details; (3) self-isolation and COVID-19 status; (4) workplace exposure characteristics; and (5) PPE ***(supplementary material)***. Free-text entries were used for gender identity (if not the same as sex at birth), ethnic background (if not within one of the listed groups), number of days of self-isolation (if greater than 14 days, with conditional limits) and an ‘Other Comments’ question. The survey covered experiences from the period 1 February 2020 to the date when each HCW participated in the study.

### Survey administration

The survey was disseminated electronically using a web link which directed potential participants to the survey form. This web link was shared on several relevant social media platforms and via e-mail. We engaged several organisations and Royal Colleges to assist with dissemination to their respective membership (a list of endorsing organisations is reported in the ***supplementary material)***.

### Definitions

Several survey response variables were grouped *a priori* to facilitate analyses. We defined a collective ‘BAME’ ethnic group as those participants who identified as ‘Asian or Asian British’, ‘Black, African, Black British or Caribbean’, ‘Mixed or multiple ethnic groups’, and ‘another ethnic, in keeping with contemporary reporting.^3,23,24^ Occupational roles were grouped into five subgroups: (1) Doctors - all doctors; (2) Dentists and dental staff - dentist, dental nurse, and dental hygienist; (3) Nurses, midwives and associated staff - healthcare assistant, maternity care worker, midwife, nurse, and nursing associate; (4) Allied Health Professionals (AHPs) - dietician, healthcare scientist (e.g. lab-based), occupational therapist, operating department practitioner, optician, paramedic, pharmacist, phlebotomist, physician associate, physiotherapist, psychologist, radiographer, speech and language therapist, technician (clinical), and therapist (Other); and (5) Other - administrative staff, domestic services, manager (care home), ‘other’, porter, senior carer (care home), support worker/assistant, and wellbeing/activity coordinator (care home). In line with Public Health England (PHE) guidance,^20^ higher risk areas were considered to be the following: COVID-19 pod/bay/ward, day case surgery unit, emergency department (ED), endoscopy unit (upper respiratory, ENT or upper GI endoscopy), intensive care (ICU)/High dependency unit (HDU), and operating theatre.

We originally included an option for “Intersex” when enquiring about sex and gender identity to support inclusivity based on published guidance.^23-25^ However, during the study, several HCWs and members of the public expressed concern regarding this approach, leading to a removal of the option for “Intersex”, leaving only “Male” and “Female” as options for sexual identity.

The primary endpoint of this study was a composite outcome of any of the following: (1) self-isolation due to COVID-19 symptoms or a positive SARS-CoV-2 test, (2) hospitalisation with suspected or confirmed COVID-19 and (3) laboratory-confirmed SARS-CoV-2 infection (via reverse transcription polymerase chain reaction or antibody testing).

### Data analysis

We report our findings according to STrengthening the Reporting of OBservational studies in Epidemiology (STROBE) guidance ***(supplementary material)**.^25^*

Statistical analyses were conducted in R Version 4.0.2 (The R Foundation for Statistical Computing, Vienna, Austria). Continuous data are reported using mean (standard deviation, SD) or median (interquartile ranges, IQR) where appropriate for measures of central tendency and spread. Categorical data are reported as numbers (percentages, %). A p-value < 0.05 was considered statistically significant. Relationships between categorical variables and outcome measures are presented as univariate odds ratios with accompanying p-values (Pearson’s Chi-square test with Yates’ continuity correction). A planned analysis of free-text entries in “Other Comments” will be reported in a separate paper.

To identify risk factors for COVID-19 amongst HCWs, we modelled the association between covariates chosen *a priori* and the COVID-19 composite endpoint using univariable and multivariable logistic regression modelling. As questions on comorbidities and tobacco smoking were optional in the survey, those participants who did not answer these questions were identified as “Prefer not to say”. For those participants who answered “No” to regular clinical contact with suspected or confirmed COVID-19 cases without adequate PPE, they were regarded as having a frequency of “None” for clinical contact without adequate PPE. Findings from the regression analysis are reported as adjusted odds ratios (OR) with 95% confidence intervals (95% CI) and accompanying p-values. Quality of the final model was assessed by the Akaike information criterion (AIC) and Area Under the Receiver Operating Characteristic curve (AUROC). Two further subgroup analyses were conducted in participants who had regular clinical contact with suspected or confirmed COVID-19 patients and in those who had regular exposure to AGPs conducted in suspected or confirmed COVID-19 patients. Code for all analyses is available on request.

### Patient and Public Involvement

As the survey was designed by HCWs, and the target population was HCWs, patient and public involvement was not sought.

## RESULTS

The study was conducted between 4-25 May 2020, and a total of 6260 participants responded, with 6152 eligible for analysis *(Figure 1)*.

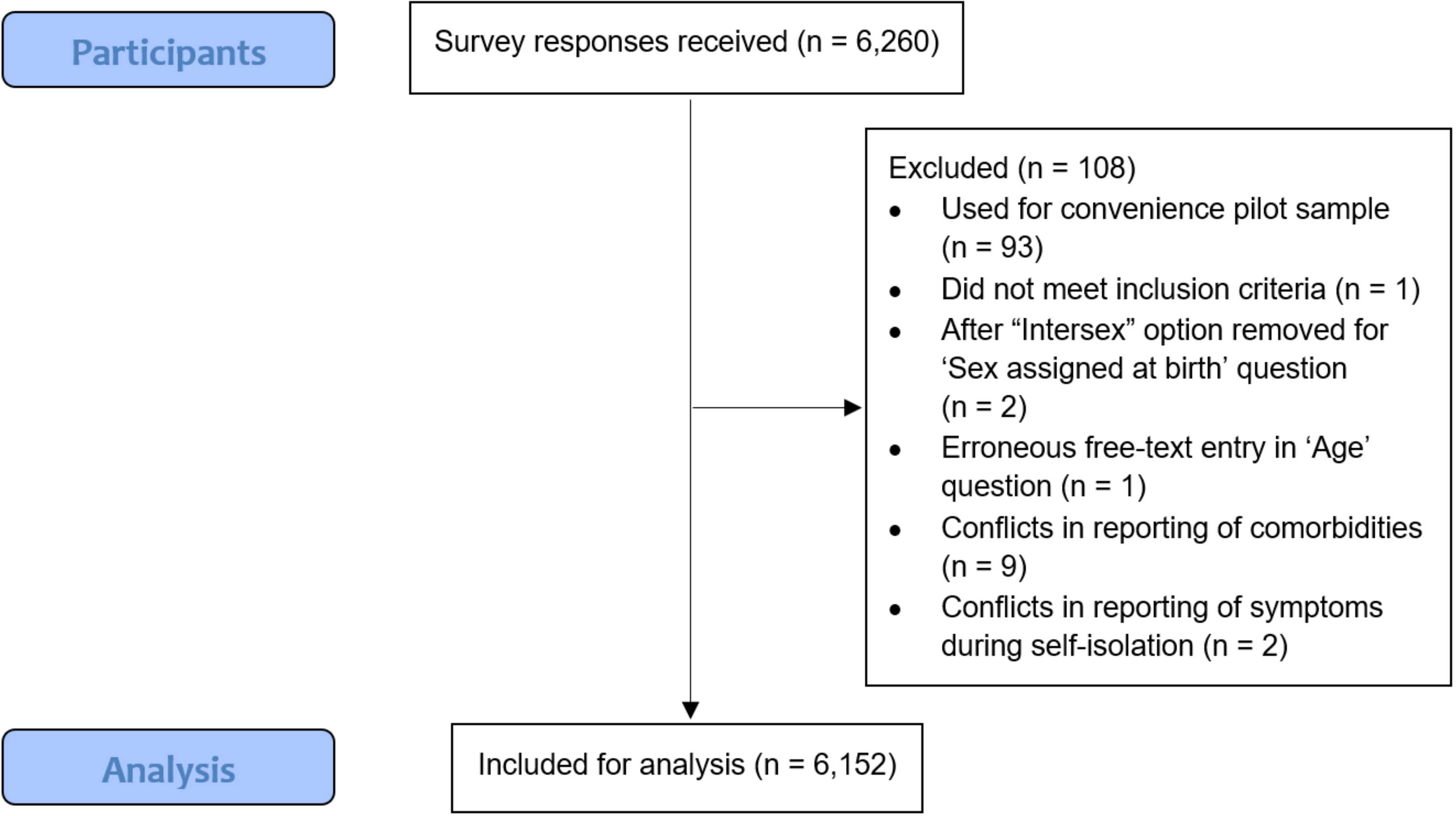

### Participant characteristics

Participant **characteristics for the sampled population are summarised in** *Table 1*.

**Table 1.**
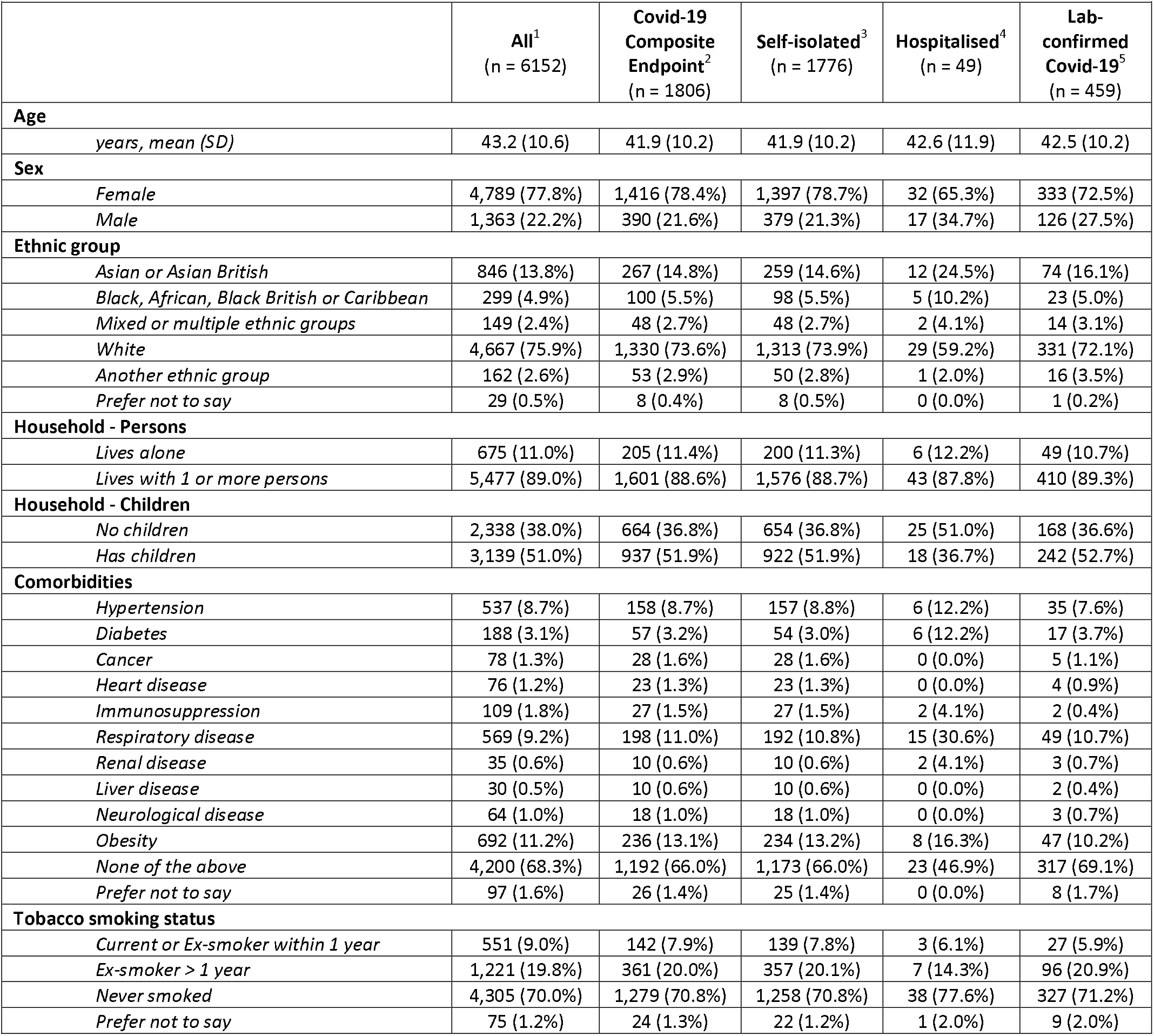
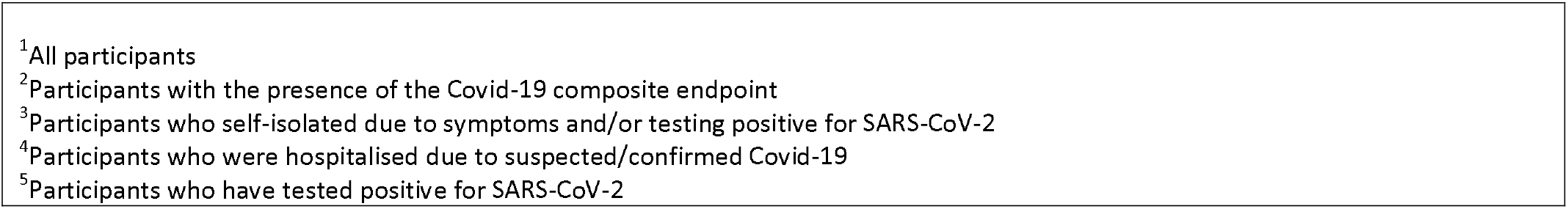
Summary of participant characteristics, stratified by covid-19 outcome. n (%) or mean (SD)

|  |  |  |  |  |  |
| --- | --- | --- | --- | --- | --- |
|  | All<sup>1</sup><br>(n = 6152) | Covid-19<br>Composite<br>Endpoint<sup>2</sup><br>(n = 1806) | Self-isolated<sup>3</sup><br>(n = 1776) | Hospitalised<sup>4</sup><br>(n = 49) | Lab-<br>confirmed<br>Covid-19<sup>5</sup><br>(n = 459) |
| **Age** |  |  |  |  |  |
| years, mean (SD) | 43.2 (10.6) | 41.9 (10.2) | 41.9 (10.2) | 42.6 (11.9) | 42.5 (10.2) |
| **Sex** |  |  |  |  |  |
| Female | 4,789 (77.8%) | 1,416 (78.4%) | 1,397 (78.7%) | 32 (65.3%) | 333 (72.5%) |
| Male | 1,363 (22.2%) | 390 (21.6%) | 379 (21.3%) | 17 (34.7%) | 126 (27.5%) |
| **Ethnic group** |  |  |  |  |  |
| Asian or Asian British | 846 (13.8%) | 267 (14.8%) | 259 (14.6%) | 12 (24.5%) | 74 (16.1%) |
| Black, African, Black British or Caribbean | 299 (4.9%) | 100 (5.5%) | 98 (5.5%) | 5 (10.2%) | 23 (5.0%) |
| Mixed or multiple ethnic groups | 149 (2.4%) | 48 (2.7%) | 48 (2.7%) | 2 (4.1%) | 14 (3.1%) |
| White | 4,667 (75.9%) | 1,330 (73.6%) | 1,313 (73.9%) | 29 (59.2%) | 331 (72.1%) |
| Another ethnic group | 162 (2.6%) | 53 (2.9%) | 50 (2.8%) | 1 (2.0%) | 16 (3.5%) |
| Prefer not to say | 29 (0.5%) | 8 (0.4%) | 8 (0.5%) | 0 (0.0%) | 1 (0.2%) |
| **Household - Persons** |  |  |  |  |  |
| Lives alone | 675 (11.0%) | 205 (11.4%) | 200 (11.3%) | 6 (12.2%) | 49 (10.7%) |
| Lives with 1 or more persons | 5,477 (89.0%) | 1,601 (88.6%) | 1,576 (88.7%) | 43 (87.8%) | 410 (89.3%) |
| **Household - Children** |  |  |  |  |  |
| No children | 2,338 (38.0%) | 664 (36.8%) | 654 (36.8%) | 25 (51.0%) | 168 (36.6%) |
| Has children | 3,139 (51.0%) | 937 (51.9%) | 922 (51.9%) | 18 (36.7%) | 242 (52.7%) |
| **Comorbidities** |  |  |  |  |  |
| Hypertension | 537 (8.7%) | 158 (8.7%) | 157 (8.8%) | 6 (12.2%) | 35 (7.6%) |
| Diabetes | 188 (3.1%) | 57 (3.2%) | 54 (3.0%) | 6 (12.2%) | 17 (3.7%) |
| Cancer | 78 (1.3%) | 28 (1.6%) | 28 (1.6%) | 0 (0.0%) | 5 (1.1%) |
| Heart disease | 76 (1.2%) | 23 (1.3%) | 23 (1.3%) | 0 (0.0%) | 4 (0.9%) |
| Immunosuppression | 109 (1.8%) | 27 (1.5%) | 27 (1.5%) | 2 (4.1%) | 2 (0.4%) |
| Respiratory disease | 569 (9.2%) | 198 (11.0%) | 192 (10.8%) | 15 (30.6%) | 49 (10.7%) |
| Renal disease | 35 (0.6%) | 10 (0.6%) | 10 (0.6%) | 2 (4.1%) | 3 (0.7%) |
| Liver disease | 30 (0.5%) | 10 (0.6%) | 10 (0.6%) | 0 (0.0%) | 2 (0.4%) |
| Neurological disease | 64 (1.0%) | 18 (1.0%) | 18 (1.0%) | 0 (0.0%) | 3 (0.7%) |
| Obesity | 692 (11.2%) | 236 (13.1%) | 234 (13.2%) | 8 (16.3%) | 47 (10.2%) |
| None of the above | 4,200 (68.3%) | 1,192 (66.0%) | 1,173 (66.0%) | 23 (46.9%) | 317 (69.1%) |
| Prefer not to say | 97 (1.6%) | 26 (1.4%) | 25 (1.4%) | 0 (0.0%) | 8 (1.7%) |
| **Tobacco smoking status** |  |  |  |  |  |
| Current or Ex-smoker within 1 year | 551 (9.0%) | 142 (7.9%) | 139 (7.8%) | 3 (6.1%) | 27 (5.9%) |
| Ex-smoker > 1 year | 1,221 (19.8%) | 361 (20.0%) | 357 (20.1%) | 7 (14.3%) | 96 (20.9%) |
| Never smoked | 4,305 (70.0%) | 1,279 (70.8%) | 1,258 (70.8%) | 38 (77.6%) | 327 (71.2%) |
| Prefer not to say | 75 (1.2%) | 24 (1.3%) | 22 (1.2%) | 1 (2.0%) | 9 (2.0%) |

<sup>1</sup>All participants
<sup>2</sup>Participants with the presence of the Covid-19 composite endpoint
<sup>3</sup>Participants who self-isolated due to symptoms and/or testing positive for SARS-CoV-2
<sup>4</sup>Participants who were hospitalised due to suspected/confirmed Covid-19
<sup>5</sup>Participants who have tested positive for SARS-CoV-2

### Work details

A total of 5518 participants were HCWs based in England (89.7%), followed by 321 (5.2%) in Scotland, 213 (3.5%) in Wales and 100 (1.6%) in Northern Ireland. Based on region, 1770 (28.8%) responses were received from Greater London, with all other regions contributing less 5.5% each. *Figure 2* shows responses stratified by main healthcare facility. Healthcare worker roles were grouped into doctors (1770 (28.8%)), nurses, midwives and associated staff (2516 (40.9%)), dentists and dental staff (198 (3.2%)), AHPs (1118 (18.2%)), and Other (550 (8.9%)). ***eTables 2*** to ***4*** summarises responses into HCW roles and grades ***(supplementary material)***.

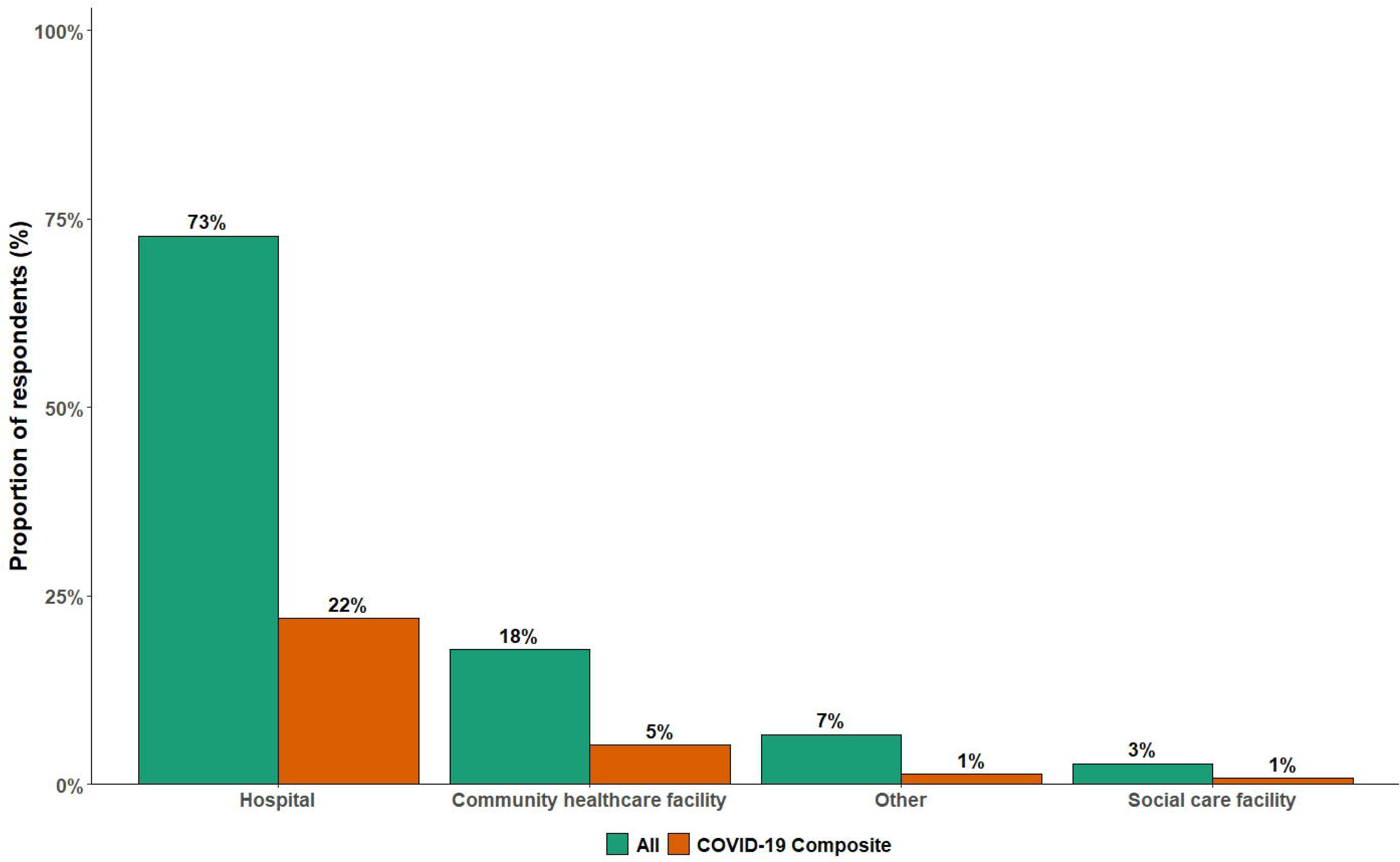

A total of 3902 (63.4%) HCWs reported regular clinical contact with suspected or confirmed COVID-19 patients. Of all participants, 2296 (37.3%) responded as having had regular exposure to AGPs performed in suspected or confirmed COVID-19 patients. Data for areas of clinical contact and the AGPs that participants were exposed to are summarised in ***eFigure 1*** and ***eFigure2***, respectively (***supplementary material)***.

### COVID-19 status

A total of 1776 participants (28.9%) self-isolated because of COVID-19 symptoms (***eFigure 3, supplementary material)*** or testing positive for SARS-CoV-2. Of those who self-isolated, 840 (47.3%) self-isolated for 1-7 days, 708 (39.9%) for 8-14 days, and 228 (12.8%) for more than 14 days. The total number of days of self-isolation in this cohort was between 11,870 and 21,158 days. The mean (SD) duration of self-isolation for participants who self-isolated for more than 14 days was 23.4 (8.8) days. In addition, 228 participants (12.8%) self-isolated more than once. Forty-nine (0.8%) participants were hospitalised for suspected or confirmed COVID-19. Responses for testing for SARS-CoV-2 revealed a total of 1407 (22.9%) participants who were tested during the period covered by the survey: 948 (15.4%) had never tested positive or were awaiting test results, 20 (0.3%) were positive on blood testing and 439 (7.1%) were positive on oral/nasal swab testing.

### Personal Protective Equipment

With regards to PPE, 4334 (70.4%) participants answered that they had received sufficient training in the use of PPE. Throughout the timeframe of interest, 1382 (22.5%) participants had been in a situation where they lacked access to items of PPE when having clinical contact with suspected or confirmed COVID-19 patients. *Figure 3* summarises the PPE items that participants reported lack of access to under these situations. Furthermore, 1306 (21.2%) participants had been in clinical contact with suspected or confirmed COVID-19 patients without adequate PPE. The top three reasons for these encounters without adequate PPE were ‘Patient not suspected/confirmed’, ‘Lack of PPE availability’, and ‘Senior instruction’ (summarised in ***eFigure 4*** of ***supplementary material)***.

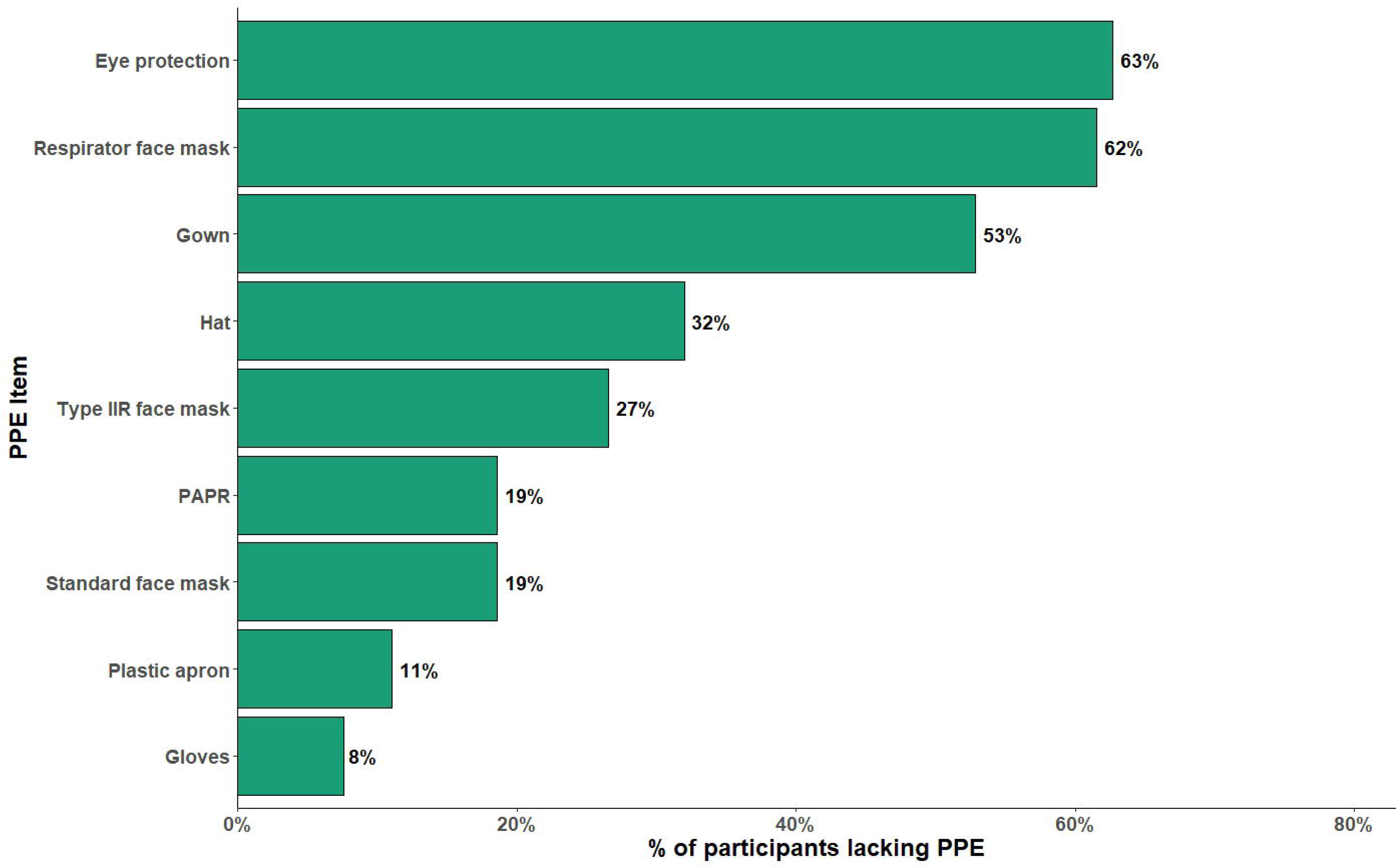

### Univariable and multivariable modelling against the COVID-19 composite

The results from univariable and multivariable analyses of covariates from the survey and the presence of the COVID-19 composite endpoint are summarised in *Table 2* and ***eTable 5 (supplementary material)***.

**Table 2.** Variables and association with the composite endpoint.

| Table 2. Variables and association with the composite endpoint |  |  |  |  |
| --- | --- | --- | --- | --- |
|  | Univariable<br>OR (95% CI) | p-value | Multivariable<br>adjusted OR (95% CI) | p-value |
| **Age** |  |  |  |  |
| *Age (per year)\** | 0.98 (0.97 – 0.99) | <0.001 | 0.98 (0.98 – 0.99) | <0.001 |
| **Sex** |  |  |  |  |
| *Female* | Ref | Ref | Ref | Ref |
| *Male* | 0.95 (0.84 – 1.09) | 0.516 | 0.92 (0.79 – 1.06) | 0.248 |
| **Ethnicity** |  |  |  |  |
| *White* | Ref | Ref | Ref | Ref |
| *BAME* | 1.19 (1.05 – 1.35) | 0.008 | 0.98 (0.85 – 1.12) | 0.747 |
| *Prefer not to say* | 0.96 (0.42 – 2.16) | 1.000 | 0.91 (0.36 – 2.06) | 0.825 |
| **Household – persons** |  |  |  |  |
| *Lives alone* | Ref | Ref | Ref | Ref |
| *Lives with 1 or more persons; no children* | 0.91 (0.75 – 1.10) | 0.344 | 0.87 (0.72 – 1.06) | 0.179 |
| *Lives with 1 or more persons; has children* | 0.98 (0.81 – 1.17) | 0.825 | 1.00 (0.83 – 1.21) | 0.962 |
| **Comorbidities** |  |  |  |  |
| *Hypertension* | 1.00 (0.83 – 1.22) | 1.000 | 1.13 (0.91 – 1.40) | 0.258 |
| *Diabetes* | 1.05 (0.76 – 1.44) | 0.831 | 1.00 (0.71 – 1.39) | 0.986 |
| *Cancer\** | 1.35 (0.85 – 2.16) | 0.249 | 1.66 (1.01 – 2.67) | 0.041 |
| *Heart disease* | 1.04 (0.64 – 1.71) | 0.962 | 1.18 (0.69 – 1.96) | 0.535 |
| *Immunosuppression* | 0.79 (0.51 – 1.22) | 0.340 | 0.83 (0.52 – 1.29) | 0.421 |
| *Respiratory disease\** | 1.32 (1.10 – 1.58) | 0.003 | 1.26 (1.04 – 1.52) | 0.015 |
| *Renal disease* | 0.96 (0.46 – 2.01) | 1.000 | 1.08 (0.49 – 2.24) | 0.833 |
| *Liver disease* | 1.20 (0.56 – 2.58) | 0.781 | 1.18 (0.51 – 2.53) | 0.686 |
| *Neurological disease* | 0.94 (0.54 – 1.63) | 0.937 | 0.88 (0.49 – 1.52) | 0.662 |
| *Obesity\** | 1.28 (1.08 – 1.52) | 0.004 | 1.31 (1.10 – 1.56) | 0.003 |
| *Prefer not to say* | 0.88 (0.56 – 1.38) | 0.657 | 0.94 (0.58 – 1.48) | 0.779 |
| **Smoking status** |  |  |  |  |
| *Never smoked* | Ref | Ref | Ref | Ref |
| *Current or Ex-smoker within 1 year\** | 0.82 (0.67 – 1.01) | 0.062 | 0.79 (0.64 – 0.98) | 0.035 |
| *Ex-smoker (more than 1 year)* | 0.99 (0.86 – 1.14) | 0.951 | 1.09 (0.94 – 1.27) | 0.238 |
| *Prefer not to say* | 1.11 (0.68 – 1.82) | 0.762 | 1.14 (0.68 – 1.87) | 0.606 |
| **Country** |  |  |  |  |
| *England* | Ref | Ref | Ref | Ref |
| *Northern Ireland\** | 0.42 (0.24 – 0.72) | 0.002 | 0.44 (0.24 – 0.75) | 0.004 |
| *Scotland* | 0.90 (0.70 – 1.16) | 0.464 | 0.95 (0.73 – 1.23) | 0.702 |
| *Wales* | 0.86 (0.63 – 1.17) | 0.379 | 1.17 (0.84 – 1.62) | 0.344 |
| **Main healthcare facility** |  |  |  |  |
| *Hospital* | Ref | Ref | Ref | Ref |
| *Community healthcare facility* | 0.94 (0.81 – 1.08) | 0.410 | 0.99 (0.84 – 1.17) | 0.940 |
| *Social care facility* | 0.95 (0.67 – 1.33) | 0.819 | 1.17 (0.81 – 1.68) | 0.399 |
| *Other* | 0.58 (0.45 – 0.74) | <0.001 | 0.82 (0.61 – 1.08) | 0.168 |
| **Role group** |  |  |  |  |
| *Nurses, midwives and associated staff* | Ref | Ref | Ref | Ref |
| *Allied health professionals\** | 0.77 (0.66 – 0.91) | 0.002 | 0.81 (0.69 – 0.96) | 0.015 |
| *Dentists and dental staff\** | 0.40 (0.27 – 0.59) | <0.001 | 0.52 (0.33 – 0.82) | 0.006 |
| *Doctors\** | 1.16 (1.02 – 1.33) | 0.025 | 1.2 (1.04 – 1.39) | 0.015 |
| *Other* | 0.84 (0.68 – 1.03) | 0.102 | 0.99 (0.78 – 1.24) | 0.915 |
| **Used public transport to travel to work \*** | 1.43 (1.26 – 1.63) | <0.001 | 1.38 (1.20 – 1.59) | <0.001 |
| **Regular clinical contact with suspected or confirmed covid-19 patients\*** | 1.52 (1.35 – 1.71) | <0.001 | 1.33 (1.15 – 1.54) | <0.001 |
| **Regular exposure to AGP(s) performed in suspected or confirmed covid-19 patients\*** | 0.97 (0.86 – 1.08) | 0.582 | 0.81 (0.71 – 0.93) | 0.004 |
| **Sufficient training in PPE use\*** | 0.79 (0.70 – 0.89) | <0.001 | 0.85 (0.75 – 0.98) | 0.023 |
| **Lacked access to PPE items for clinical contact** | 1.74 (1.53 – 1.97) | <0.001 | 1.28 (1.09 – 1.51) | 0.002 |
| **with suspected or confirmed covid-19 patients\*** |  |  |  |  |
| **Clinical contact without adequate PPE** |  |  |  |  |
| *Never* | Ref | Ref | Ref | Ref |
| *Rarely* | 1.35 (1.13 – 1.62) | 0.001 | 1.06 (0.87 – 1.29) | 0.547 |
| *Sometimes\** | 2.32 (1.92 – 2.79) | <0.001 | 1.7 (1.37 – 2.10) | <0.001 |
| *Often\** | 2.56 (1.83 – 3.58) | <0.001 | 1.84 (1.28 – 2.63) | 0.001 |
| *Always\** | 3.65 (2.18 – 6.10) | <0.001 | 2.93 (1.72 – 5.06) | <0.001 |
| **Reused disposable PPE** | 1.21 (1.07 – 1.37) | 0.002 | 0.98 (0.86 – 1.13) | 0.821 |
| **Used improvised PPE\*** | 1.10 (0.94 – 1.28) | 0.271 | 0.81 (0.68 – 0.97) | 0.020 |
| Univariate and multivariate odds ratio (OR), 95% confidence intervals (95%CI). |  |  |  |  |
| * *p*-value <0.05 for multivariable model. |  |  |  |  |
| Abbreviations: Ref = Reference value, BAME = Black, Asian and Minority Ethnic, AGP = Aerosol-Generating Procedures, PPE = Personal Protective Equipment. |  |  |  |  |

No difference in the presence of the COVID-19 composite endpoint was seen between different ethnic groups. This persisted with constituent ethnic groups replacing the collective BAME group: Asian or Asian British (adjusted OR 0.96 (0.81 - 1.14), p-value = 0.643), Black, African, Black British or Caribbean (adjusted OR 1.03 (0.79 - 1.33), p-value = 0.845), Mixed or Multiple ethnic groups (adjusted OR 0.99 (0.69 - 1.40), p-value = 0.937), and Another ethnic group (adjusted OR 0.96 (0.67 - 1.36), p-value = 0.828) ***(eTable 6, supplementary material)***.

Being a current or ex-smoker (within 1 year) was associated with a significant decrease in odds for the COVID-19 composite endpoint compared to participants who never smoked. To assess if this effect was due to collider bias following adjustments for comorbidities in our model, an additional model was constructed with comorbidities removed ***(eTable*** **7**, ***supplementary material)***. However, in this model, being a current or ex-smoker (within 1 year) still had reduced odds for the presence of the composite endpoint (adjusted OR 0.79 (0.64 - 0.98), p-value = 0.034).

In a subgroup analysis of those who had regular clinical contact with suspected or confirmed COVID-19 patients (3902 (63%)), participants working in higher risk areas as defined by PHE^15^ made up 81.7% of this subgroup. Working in an inpatient clinic area was associated with a significant increased risk of reporting the primary endpoint (adjusted OR 1.41 (1.01 - 1.97), p = 0.043). The following areas were associated with a significant decreased risk: home visits (adjusted OR 0.68 (0.47-0.98), p = 0.040), ICU/HDU (adjusted OR 0.78 (0.65 - 0.94), p = 0.007), operating theatre (adjusted OR 0.71 (0.57 - 0.87), p = 0.001), radiology (adjusted OR 0.62 (0.42 - 0.91), p = 0.016) and other areas (adjusted OR 0.69 (0.48 - 0.98), p = 0.044). ***eTable 8 (supplementary material)*** summarises the model for this subgroup analysis.

In terms of exposure to AGPs, a separate subgroup analysis of participants who had been regularly exposed to AGPs used in suspected or confirmed COVID-19 patients (2296 (37.3%)) showed that 95.8% of this cohort were exposed to procedures considered by PHE to be AGPs^18^ (i.e. this excludes ‘CPR’, ‘nebulisers’ and ‘other’ as AGPs during the study period). In this subgroup, no particular AGP was associated with a significant change in risk on multivariate analysis ***(eTable 9, supplementary material)***.

## DISCUSSION

### Principal Findings

We describe the characteristics of a sample of UK-based HCWs working during the COVID-19 pandemic and relate their experiences to the development of COVID-19 infection-related outcomes. The overall prevalence of the primary composite endpoint amongst HCWs was 29.4% over the period from 1^st^ February to 25^th^ May 2020. A number of risk factors were explored using regression modelling of the survey responses. Additionally, we report a substantial number of working days lost from self-isolation due to symptoms and estimate between 11,800 and 21,100 working days lost during the study period, translating to between 71 to 127 working days lost per 1000 working days (assuming a 40-hour work week per HCW). We also report that 22.5% of participants had encountered a situation where they lacked PPE items and identified a variety of PPE items that were not available.

### Comparison with other studies

The prevalence of suspected or confirmed COVID-19 amongst HCWs is higher in our sampled population compared to other sources.^5,6,11-13^ Previous estimates have ranged from as low as 1.73% through a population survey-based approach to as high as between 7.7 - 18% via testing of healthcare staff.^6,12,13^ A study amongst HCWs involved in tracheal intubation using a similar primary endpoint reported an overall incidence of 10.7% over a median follow-up period of 32 days.^5^ The use of a composite endpoint facilitated capture of outcomes from individuals who were plausibly at risk of testing positive for SARS-CoV-2, but who were never tested. Indeed, 77.2% of our total sampled population (n = 4745) had never been tested. During the time period, HCWs had to self-isolate based on clinical symptoms alone due to lack of mass testing.^26^ Drawing definitive prevalence conclusions from the data reported herein is challenging due to the self-reported nature of study conduct, but the magnitude of the reported prevalence cannot be ignored.

Our data did not suggest any difference between White and BAME groups within the HCW population for developing the COVID-19 composite endpoint, after adjusting for comorbidities (including obesity). However, amongst the hospitalised group, there was a higher proportion of BAME HCWs compared with the total sampled population (40.8% vs. 23.7%), particularly ‘Asian or Asian British’, and ‘Black, African, Black British or Caribbean’. Increased focus on the BAME community has resulted from findings of more severe COVID-19 infection amongst individuals of BAME origin.^3,4,23^ PHE have previously reported on the disparities in risks and outcomes for COVID-19 infection, identifying a higher prevalence of positive tests forSARS-CoV-2 and more severe disease amongst BAME groups within the UK, though the effects of occupation and comorbidities (including obesity) were unaccounted for.^27,28^ Thus, despite similar susceptibilities to the disease, it remains the case that HCWs from BAME origins may be at risk of more severe disease and death.

Adequate training and correct use of PPE (particularly during donning and doffing) are important in reducing the risk of transmission of respiratory infectious disease from patient to HCWs ‘ ‘ and this was reflected in our results. This may also explain our finding that HCWs exposed to regular AGPs in suspected or confirmed COVD-19 patients were less likely to have the presence of the primary endpoint. Given the importance of PPE use to protect against viral transmission,^29,31,32^ it is unsurprising that participants who lacked access to PPE items, and those who were more frequently exposed to suspected or confirmed cases of COVID-19 without adequate PPE had a higher risk of the presence of the COVID-19 composite endpoint. That nearly a quarter of UK HCWs reported being in such a situation is notable.

### Strengths and Limitations

Strengths of our study include a relatively large sample size and the inclusion of HCWs from all backgrounds and work environments to facilitate risk comparisons using a standardised survey. We captured granular information that has otherwise been poorly reported in prevalence studies in HCWs. For example, medical history and details regarding the use, or lack thereof, of PPE have not been elsewhere reported. We did not limit our recruitment to primary or secondary care, and thus the associations we report are generalisable across a wide range of healthcare settings.

Several limitations need to be addressed. First, data were gathered using a survey-based approach which risks selection and recall bias. We also could not capture data from HCWs who died from COVID-19 infection, or those who were too ill to respond. However, our methodology allowed us to rapidly capture both objective and subjective granular data from a large number of participants. Second, we were unable to determine a denominator to quantify a response rate for this observational study. Third, the use of a composite outcome to detect suspected or confirmed COVID-19 infection in HCWs may have resulted in an overestimation of prevalence. However, this definition is in keeping with that used in other studies^5^ and internationally.^7^ Availability of testing for HCWs was also limited during early phases of the pandemic, thus clinical diagnoses were often relied upon. On the other hand, data have estimated that 7% of HCWs are asymptomatic seroconverters,^11,33^ and thus our data could potentially represent an underestimation of COVID-19 transmission. Fourth, we sought some subjective data, although this was a pragmatic decision to maximise detail in responses. Fifth, several changes to national guidance and policies were made throughout the study period^15,37^ which may confound responses regarding PPE. Finally, all data herein are subjective and represent hypothesis-generating associations in the responding participants. Objective analyses are required to determine the findings we reported.

### Conclusions and Implications

We found a reported prevalence estimate of suspected or confirmed COVID-19 infection of nearly a third, based on a COVID-19 composite endpoint, amongst HCWs within the UK. We also present several risk factors associated with reporting of this endpoint, lack of PPE being an important consideration. As a consequence of self-isolation, between 11,000 to 21,000 days of clinical service was lost. In future, important considerations for policymakers are to ensure adequate PPE supplies to all HCWs in preparation for a potential surge in COVID-19 cases and accessible, rapid, accurate testing strategies to enable improved healthcare workforce planning.

## Data Availability

Additional data is available upon request and agreement with the corresponding author, KE, by emailing.

## ACKNOWLEDGEMENTS

The authors would like to thank all of the 6260 healthcare workers who participated in the healthcareCOVID study. In addition, we would like to thank Steve Palmer and Knack (Philadelphia, USA) for providing assistance with the web-based software, database and server space used for the healthcareCOVID study.

## Contributorship statement

JK, and KE conceptualised the study. JK, RP, CM, DJNW, CM, IA, and KE contributed to study design, survey design and web application development. All authors were involved in promotion and administration of the survey. JK, and DJNW performed the statistical analyses. JK, RP, and DJNW drafted the manuscript. All authors contributed to revisions toward the final manuscript. KE is the corresponding author, and attests that all listed authors meet authorship criteria and that no others meeting the criteria have been omitted.

## Guarantor information

JK is the guarantor.

## Data sharing statement

Additional data is available upon request and agreement with the corresponding author, KE, by e-mailing.

## Transparency declaration

JK affirms that this manuscript is an honest, accurate, and transparent account of the study being reported; that no important aspects of the study have been omitted; and that any discrepancies from the study as planned (and, if relevant, registered) have been explained.

## Dissemination declaration

Summary reports of the study have been provided to relevant stakeholder organisations and to study participants who wished to be informed (as part of their responses to the survey).

